# Patients with chronic cluster headache may show reduced activity energy expenditure on ambulatory wrist actigraphy recordings during daytime attacks

**DOI:** 10.1101/2023.10.05.23296527

**Authors:** Nicolas Vandenbussche, Jonas Van Der Donckt, Mathias De Brouwer, Bram Steenwinckel, Marija Stojchevska, Femke Ongenae, Sofie Van Hoecke, Koen Paemeleire

## Abstract

**Objective:** To investigate the changes in activity energy expenditure throughout daytime cluster headache attacks in patients with chronic cluster headache, and to evaluate the usefulness of actigraphy as a digital biomarker of cluster headache attacks.

**Background:** Cluster headache is a primary headache disorder characterized by attacks of severe to very severe unilateral head pain with ipsilateral cranial autonomic symptoms and/or a sense of restlessness or agitation. We hypothesized increased activity energy expenditure from hyperactivity during attacks measured by actigraphy.

**Methods:** An observational study including patients with chronic cluster headache was conducted. During 21 days, patients wore an actigraphy device on the non-dominant wrist and recorded cluster headache attack-related data in a dedicated smartphone application. Accelerometer data was used for the calculation of activity energy expenditure before and during daytime cluster headache attacks that occurred in ambulatory settings, and without restrictions on acute and preventive headache treatment. We compared the activity and movements during the pre-ictal, ictal, and postictal phases with data from wrist-worn actigraphy with time-concordant intervals during non-headache periods.

**Results:** Four patients provided 34 attacks, of which 15 attacks met the eligibility criteria for further analysis. In contrast with the initial hypothesis of increased energy expenditure during cluster headache attacks, a decrease in movement was observed during the pre-ictal phase (30 min before onset to onset) and during the headache phase. A significant decrease (p<0.01) in the proportion of high-intensity movement during headache attacks, of which the majority were oxygen-treated, was observed. This trend was less present for low-intensity movements.

**Conclusion:** The results of our analysis may shift the initial hypothesis for future research towards a decrease in activity energy expenditure during the pre-ictal and headache phase of daytime attacks in patients with chronic cluster headache under acute and preventive treatment in ambulatory settings.

## Introduction

Cluster headache (CH) is a primary headache disorder characterized by attacks of severe to very severe unilateral head pain lasting 15 to 180 minutes when untreated ^1^. The International Classification of Headache Disorder Third Edition (ICHD-3) criteria for CH furthermore require that attacks are associated with cranial autonomic symptoms, e.g. lacrimation, and/or a sense of restlessness or agitation ^1^. Pre-ictal symptoms were only recently studied in detail and are in fact very common; they include both local and general symptoms ^2^. Patients are diagnosed with episodic cluster headache (ECH) if they have periods of CH attacks that are separated by pain-free periods lasting at least 3 months, while the diagnosis of chronic cluster headache (CCH) is given when CH attacks occur for at least one year without a remission period or with remissions lasting less than 3 months ^3^.

Patient descriptions and clinical observations indicate that hyperactivity and/or increased movement may be an associated symptom during CH attacks. Ekbom was the first to report that patients are unable to sit still during a CH attack, this in contrast to migraine patients experiencing increased pain with head movements ^4^. Blau asked patients to act out their behaviors during CH attacks and these included pacing while clutching the head, sitting and rocking backwards and forwards while holding the head, pressing the affected eye or temple, and hitting the forehead ^5^. Even violent, destructive behavior and self-inflicted injuries have been described ^4–7^. These behaviors seemed to be related to the severity of the pain ^8^. More recently CH pain has been conceptualized as exteroceptive pain with associated fight-flight response including motor restlessness and agitation, in contrast to migraine pain described as visceral pain ^9^.

Actigraphy is a non-invasive method that utilizes continuous monitoring sensors worn on the body to objectively measure movement and physical activity ^10^. Therefore, actigraphy may provide insights into patient behaviors during the pre-ictal, ictal and postictal phases of CH attacks. The primary objective of our research is to investigate the differences in activity energy expenditure (AEE) during CH attacks using a wrist-worn actigraphy device in unconstrained ambulatory environments ^11^. In line with the existing medical literature, we hypothesize that AEE measured by wrist-worn actigraphy increases during attacks. A secondary objective of the study is to examine AEE changes in the pre-ictal phase. By hypothesizing that restlessness and agitation can be measured using actigraphy, this technique holds promise as a digital biomarker for identifying CH attacks.

## Methods

### Participants

Patients with a diagnosis of CCH (ICHD-3 diagnosis 3.1.2), recruited within the tertiary headache clinic of the Ghent University Hospital, and not suffering from any other headache syndrome (except for infrequent tension-type headache), were included in the study.

Inclusion criteria were: age between 18 and 65 years, at least 5 attacks per week expected, no significant medical comorbidity interfering with movement and activity, no drug or alcohol abuse, and no use of beta-blockers.

All participants had access to their habitual attack treatment of subcutaneous sumatriptan 6mg or high flow oxygen at 12-15 liters per minute via a non-rebreathing mask and other acute treatments at their own discretion. There were no restrictions on the use of preventive treatments for CH for the duration of the study.

### Wrist-worn actigraphy sensor and smartphone application

The Empatica E4® device was used for the actigraphy recordings on the non-dominant wrist ^12^. This device is a CE-certified medical-grade wearable that offers physiological data acquisition. It has an internal memory that can store up to 60 hours of data, but in this study the data was sent in real-time over a Bluetooth Low Energy connection to a smartphone that streamed the data to our cloud. Empatica E4® devices have an onboard microelectromechanical system type 3-axis accelerometer that measures continuous gravitational force (g) applied to each of the three spatial dimensions (x, y, and z). The scale is set to +-2g. These accelerometer signals are collected at 32 Hz for the three dimensions.

Participants manually documented headache attacks using a dedicated headache smartphone application developed for this study (Figure 1). The recorded attack data included the time of onset (hour and minutes) and end (hour and minutes) of each CH attack, the intensity of the attack measured on a 5-point Likert Scale ranging from no pain to very severe pain, the use of attack medication and the effectiveness of those treatments (Part b of Figure 1) ^13,14^.

**Figure 1:**
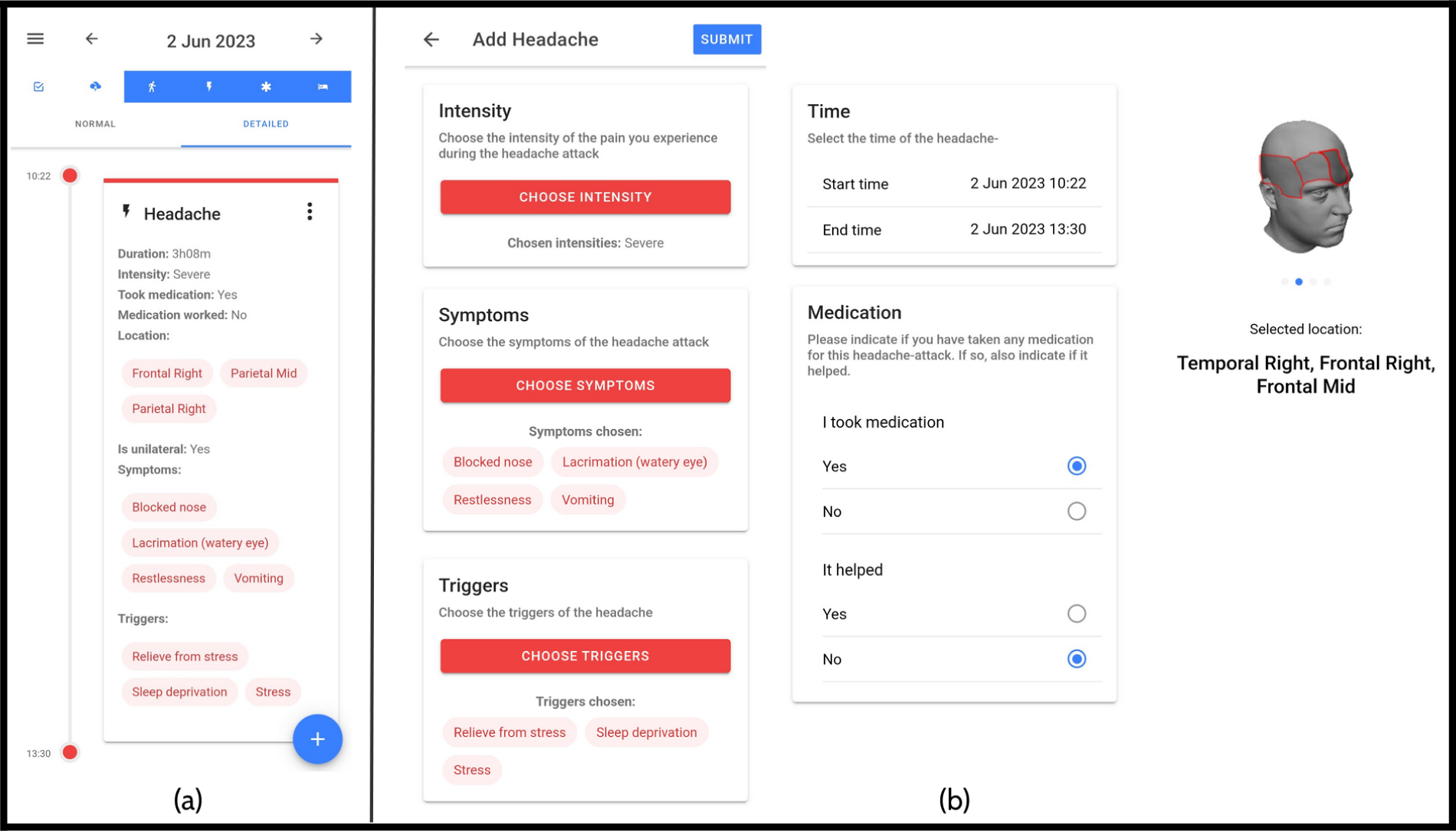
Illustration of mBrain app timeline view (a) and corresponding headache registration (b).

### Study design

The study consisted of a baseline study visit and a final study visit 21 days later. During the study period, participants were instructed to wear the device as much as possible (daytime and nighttime) which ensured consistency in data collection and minimized disruption to daily routines. They were also advised to charge the sensor device at least once a day in the evening. Participants were asked to perform their regular daily activities during the measurement period. However, they were asked to take off the Empatica E4 device when engaging in activities involving water (e.g. showering, swimming), heat (e.g. barbecuing) or cold (e.g. working in freezers).

### Absolute Activity Index as a marker for Activity Energy Expenditure

From the time series data of g-force measurements, the absolute activity index (AI^ABS^) is calculated and used as the metric of AEE ^15^. As shown in Figure 2 (a), the AI^ABS^ is formulated as the square root of the mean variance of the raw accelerometer signals, with the variance computed over a fixed period, H. The AI^ABS^ exhibits a robust correlation with activity intensity ^15^. Empirical analysis using the Empatica device revealed that the systematic noise variance, *σ_i,_*was negligible, and therefore *σ_i_*was set to 0 for the calculation of the AI^ABS^, leading to a simplification of the AI^ABS^ formula. In alignment with Bai et al., the AI^ABS^ is computed over a time-period H of 1 second, corresponding to 32 sampling points ^15^. Afterward, also in accordance with Bai et al., these second-by-second AI^ABS^ values are aggregated to a per-minute AI, by averaging the 1-second AI^ABS^ values within each 1-minute period (Figure 2 part b). For time intervals of interest, such as the CH headache attack period, the AI^ABS^ values can be represented by a distribution. Considering the non-normal distribution of AI^ABS^ over these intervals, as depicted in Figure 2 (c), we employ a range of quantile-based features for analysis. Specifically, we calculate the median AI^ABS^ (i.e., 50th percentile), as well as the 25th, 75th, and 90th percentiles. We included percentile 90 to examine movement associated with higher energy expenditure, in alignment with our hypothesis.

**Figure 2:**
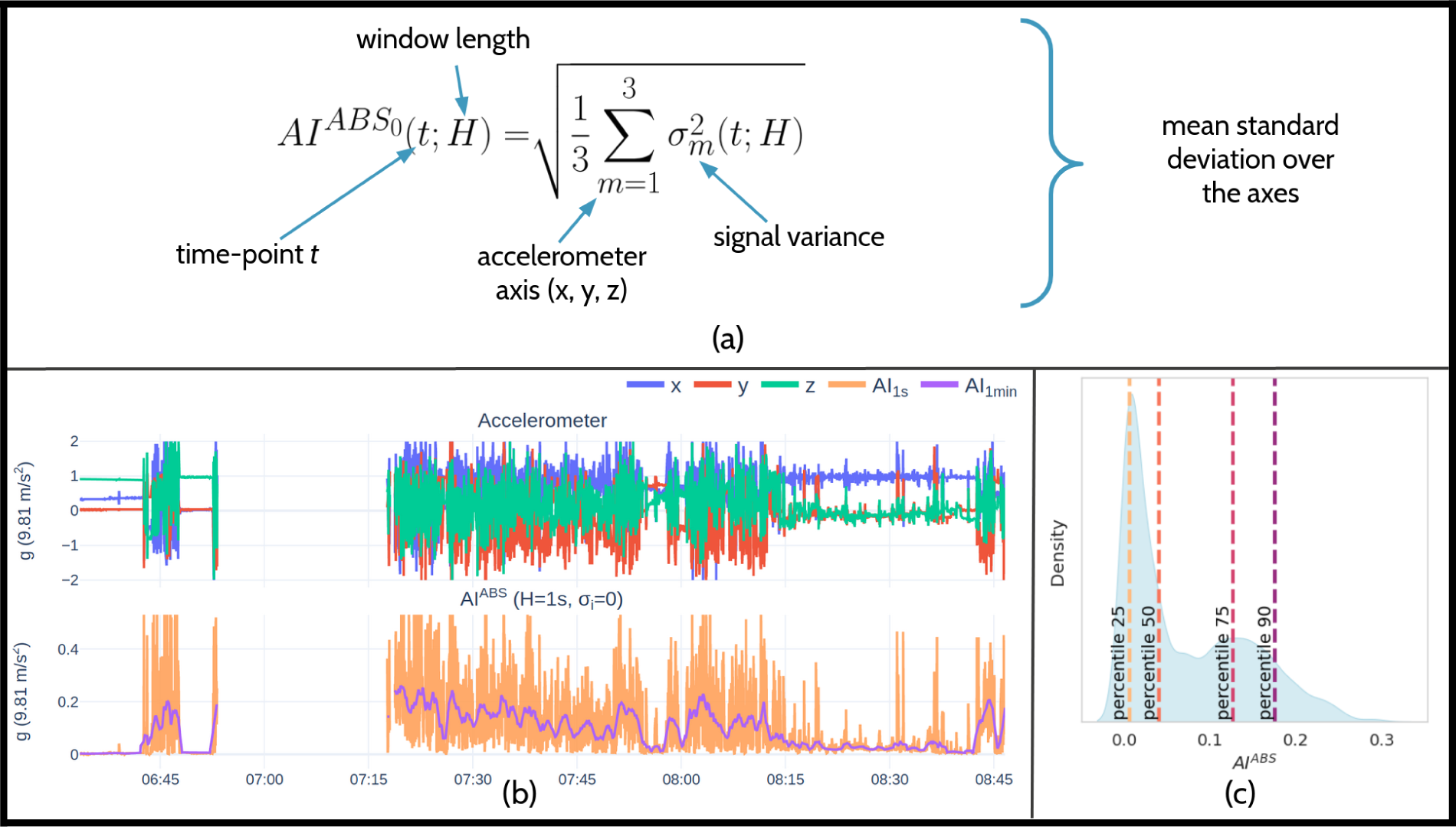
Illustration of AI^ABS^ computation and its distribution properties. To calculate the AI^ABS^, the simplified equation given in (a) was used, which assumes a systematic noise-variance *σ_i_* of zero. Subplot (b) portrays the 1-minute aggregated AI^ABS^ for an Empatica accelerometer excerpt. Subfigure (c) demonstrates the distribution and percentiles of the AI^ABS^ of (b). *Abbreviations*: H = variance window length, t = time point, σ_m_ = signal variance of wrist acceleration over axis x, y or z.

### Eligibility criteria for analysis of CH attacks and corresponding non-CH periods

To be eligible for analysis, CH attacks had to fulfill the following criteria. The attacks must have occurred during daytime periods without overlap between the headache period and nighttime (between 11:00 PM and 7:30 AM). Additionally, a minimum data ratio of 75% during headache attacks was utilized to ensure a reliable analysis of actigraphy data (i.e., no more than 25% of data missing during the attack).

For comparison purposes, periods without CH attacks from the same participant were used, subject to the following conditions. Non-CH periods had to be at least 24 hours distant from the start and end of CH attacks, and a time range equal to the CH attack was used on the same type of day, either weekdays or during weekends. In cases when multiple eligible non-CH periods were identified, these periods were accumulated to construct a single non-CH AI^ABS^ distribution to be paired against the corresponding CH attack. Data from Belgian holidays were excluded from the analysis.

### Analysis and statistics

Demographic data and participant-specific information (i.e., age, sex, diagnosis, age of onset and treatment regimens related to CH) are provided in a descriptive manner as proportions and means with standard deviations (SD). Recorded CH attacks are described with mean and SD of attack duration (in minutes), mean and SD of attack intensity, the proportion of attacks treated with acute therapy, and proportion of acute-treated attacks which were successfully managed.

The first analysis assesses AEE during attacks by using each CH headache period as time range. Specifically, we calculated AI^ABS^ distributions for eligible single CH attacks and their corresponding non-CH data. Percentile AI^ABS^ differences (ΔAI^ABS^) for single attacks were determined by subtracting the CH attack percentile AI^ABS^ value from the non-CH data’s percentile value. Furthermore, this analysis also assesses the impact of acute treatment type, which is represented via a color hue.

The second analysis examined AEE distributions for specific time intervals relative to headache onset, including 3 to 1 hour before onset, 1 hour to 30 minutes before onset, 30 minutes to onset, onset to 30 minutes after, 30 minutes to 1 hour after, 1-2 hours after, and 2-3 hours after. Percentile AI^ABS^ interval differences were calculated by subtracting the CH attack interval percentiles from the corresponding non-CH interval percentiles. The number of events may vary for each interval, as the ≥75% data ratio must be fulfilled for both CH and non-CH intervals.

Given the high likelihood of non-normality in the separate AI^ABS^ interval distributions as depicted by Figure 2 (c), the Wilcoxon signed-rank test was employed to evaluate the statistical significance of AI^ABS^ percentile pairs for CH and their corresponding non-CH intervals. A two-tailed test was utilized for each of those analyses. Afterward, all results were subjected to Bonferroni correction for multiple testing, therefore only p-values corrected for multiple testing are presented.

Due to the exploratory nature of our pilot study and the lack of previous data, no formal sample size calculation was performed before the onset of the study.

All data processing and transformations were conducted via Python 3.8. Statistical testing was performed using the SciPy package ^16^. For exploratory data analysis of the raw wearable data, the Plotly-Resampler tool was utilized ^17^. Via this exploratory analysis, we assessed whether the time-ranges of the headache periods overlapped with typical sleep periods. Using the 11PM-7:30AM nighttime filter, we observed that none of the remaining daytime CH intervals fell within the patient’s typical sleep periods. During the exploratory analysis of the raw wearable data, off-body periods were observed, which refer to intervals where wearable data was present despite the device not being worn ^18^. To address this issue, an on-body detection algorithm was utilized to identify and exclude off-body periods ^19^. This processing step was performed prior to computing the interval data ratios and the AI^ABS^. In order to efficiently compute the AI^ABS^, numPy functions were leveraged through the tsflex library ^20,21^.

### Ethics

This study was approved by the Committee for Medical Ethics of the Ghent University Hospital (internal ID BC-07403, approved June 12^th^ 2020). Patients were fully informed on all the aspects of the study (duration, procedures, study visit, etc.) and gave written informed consent at the beginning of the study. Participants received a pseudonymized code throughout the study. Only physician-researchers had the key to decode the participant if required.

## Results

### Participants and CH attacks

Four male participants with CCH were included in the analysis (Table 1). All participants had access to high-flow oxygen and/or sumatriptan 6mg SC injection for the acute treatment of CH attacks. All participants used preventive treatment with verapamil, and two participants used melatonin before bedtime.

**Table 1:** Descriptive characteristics of participants (N=4).

| Patient | Diagnosis | Age | Sex | Age at onset CH | Acute treatment | Preventive treatment | Total CH attacks / Daytime / ... &<br>>=75% data / ... &<br>eligible non-CH data |
| --- | --- | --- | --- | --- | --- | --- | --- |
| 1 | CCH | 50s | Male | 35-40 | Oxygen, Sumatriptan SC 6 mg | Verapamil 240 mg daily dose | 5 / 3 / 3 / 3 |
| 2 | CCH | 60s | Male | 60-65 | Oxygen, Sumatriptan SC 6 mg | Verapamil 480 mg daily dose | 14 / 9 / 6 / 6 |
| 3 | CCH | 40s | Male | 35-40 | Oxygen, Sumatriptan SC 6 mg | Verapamil 720 mg daily dose, Melatonin 3 mg before bedtime | 7 / 1 / 1 / 0 |
| 4 | CCH | 20s | Male | 14-18 | Oxygen, Sumatriptan SC 6 mg, Zolmitriptan nasal spray 5mg, Paracetamol 250mg/Acetylsalicylic acid 250mg/Caffeine 65mg | Verapamil 640 mg daily dose, Melatonin 5mg before bedtime | 8 / 8 / 6 / 6 |

Abbreviations: CH = Cluster Headache; CCH = chronic cluster headache; mg = milligram; SC = subcutaneous.

In total, 34 CH attacks were recorded. Fifteen of these attacks from 3 participants met the daytime and data ratio requirements, while also having eligible non-headache period(s) and were used for further analysis (7 out of 7 attacks from 1 participant could not be used due to 6 nighttime attacks and 1 attack not having an eligible non-CH interval). The mean duration of the analyzed attacks was 38 minutes (SD 28 minutes). Thirteen out of the 15 attacks (87%) were treated with acute therapy: 8 with high-flow oxygen, 2 with combination analgesics paracetamol 250mg/acetylsalicylic acid 250mg/caffeine 65mg, 1 with combination high-flow oxygen and zolmitriptan 5mg nasal spray, and 2 with non-specified treatments. Twelve of these thirteen attacks were found to be treated effectively by the participants (92%) (Table 2). The mean amount of data ratio per individual attack was 98.0% (SD 6%). For every analyzed CH attack, the corresponding volume of eligible non-CH data was equal to or higher than the duration of the CH attack itself.

**Table 2.** Descriptive analysis of all cluster headache attacks registered during study.

| Description | N | Mean duration (SD), minutes | Mean intensity (SD) * | Acute treatment use (%) / (n/N) | Acute treatment effectiveness (%) / (n/N) |
| --- | --- | --- | --- | --- | --- |
| All headaches | 34 | 35 (22) | 2.9 (0.7) | 82% (28/34) | 96% (27/28) |
| Nighttime headaches | 13 | 32 (21) | 3.3 (0.7) | 69% (9/13) | 100% (9/9) |
| Daytime headaches | 21 | 37 (23) | 2.6 (0.7) | 90% (19/21) | 95% (18/19) |
| Daytime headaches w/ ≥75% data during attack | 16 | 38 (25) | 2.6 (0.7) | 88% (14/16) | 93% (13/14) |
| Daytime headaches w/ ≥75% data during attack & eligible non CH period | 15 | 38 (26) | 2.5 (0.6) | 87% (13/15) | 92% (12/13) |

Note: \*: Intensity levels: 1 = no pain, 2 = light pain, 3 = moderate pain, 4 = severe pain, 5 = very severe pain.
Abbreviations: n, N = number; SD = standard deviation.

### Activity Energy Expenditure during CH attacks and by acute treatment type

Percentile ΔAI^ABS^ values were calculated for the headache periods of the 15 qualifying attacks, as illustrated in Figure 3. When compared to the corresponding non-headache periods, the AI^ABS^ values during CH attacks show a negative ΔAI^ABS^ trend, indicating a reduction in AEE. This reduction is also represented by a significantly lower 75th percentile (p=0.007) and 90th percentile (p=0.0014). No significant differences in AI^ABS^ values were found for the 25th percentile (p=0.205) and median (p=0.054) between CH attacks and non-headache periods). Figure 3 also uses color hues to denote the acute treatment type used during a single CH attack. For all four percentiles, all ΔAI^ABS^ values are negative for the CH attacks that were treated with oxygen, indicating no increases in AEE for all of the 9 oxygen-treated CH attacks. Appendix 3 presents the statistical significance results exclusively for the oxygen treatment group (N=9).

**Figure 3:**
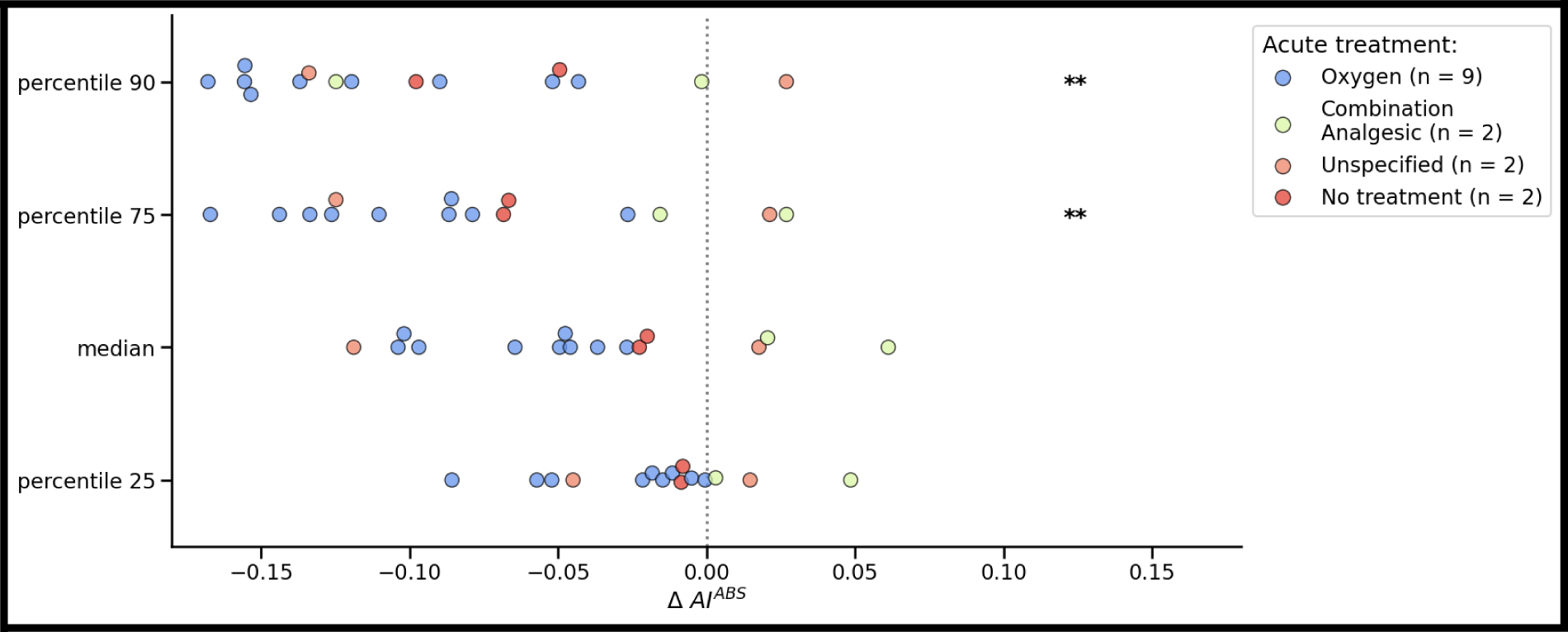
Differences in percentile AI^ABS^ (ΔAI^ABS^) during individual attacks and acute treatment type. ΔAI^ABS^ is determined by subtracting the AI^ABS^ percentile value of each CH attack from its corresponding eligible non-CH value (N=15). Each dot represents the difference for a single CH attack. The acute treatment is represented via a color hue. The treatment types include: No treatment (N=2), Unspecified (N=2), Oxygen (N=9), and Combination Analgesic (N=2). An “Unspecified” treatment indicates that the participant reported using medication during the headache event, but no specific medication event was reported afterward. *Abbreviations*: AI^ABS^ = absolute activity index, ΔAI^ABS^ = difference between absolute activity indices. Levels of statistical significance (after Bonferroni correction for multiple testing): * p < 0.05; ** p < 0.01; *** p < 0.001.

### Temporal patterns of Activity Energy Expenditure before and during CH attacks

The decrease of intense movements during headaches, resulting in negative ΔAI^ABS^ values, is observed in Figure 3. This decrease is also reflected in Figure 4 by the negative yet non-significant ΔAI^ABS^ trend of the “onset to 30 min” interval (N=14) for the 75th (p=0.215) and 90th percentile (p=0.098). Additionally, Figure 4 shows that this negative ΔAI^ABS^ trend is already observed during the pre-ictal phase, specifically for the “-1h to -30min” (N=12) and “-30min to onset” (N=11) intervals at the 75th and 90th percentiles. The “30 min to 1h’’ (N=13) interval also exhibits a decreasing, but non-significant, trend. This negative ΔAI^ABS^ trend persists up to 3 hours after onset. No statistically significant differences were observed for any of the interval percentiles, which could be attributed to the reduced sample size of the intervals. Analysis for the postictal phase can be found in Appendix 5.

**Figure 4:**
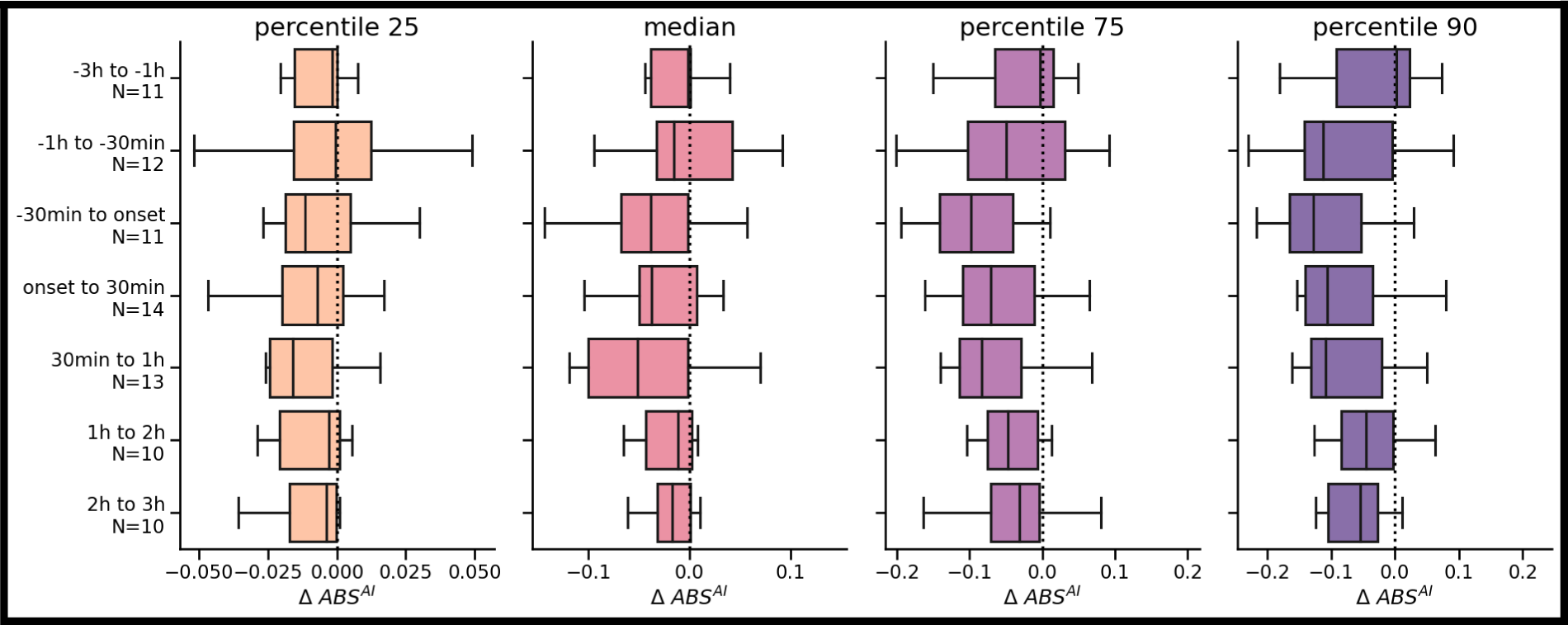
**Percentile AI^ABS^ differences (**Δ**AI^ABS^ distributions) for time ranges relative to the headache onsets.** The y-axis labels indicate the number of attacks considered, which varies due to the >=75% data ratio requirement for both CH and non-CH intervals. *Abbreviations*: AI^ABS^ = absolute activity index, ΔAI^ABS^ = difference between absolute activity indices, CH = cluster headache, h = hour, N = number. Levels of statistical significance (after Bonferroni correction for multiple testing): * p < 0.05; ** p < 0.01; *** p < 0.001.

## Discussion

To our knowledge, this is the first objective, real-world study utilizing wrist-worn actigraphy to measure the AEE before and during CH attacks in patients with CCH during daytime in the ambulatory setting. Actigraphy in the field of CH has only been used in sleep studies so far ^22,23^. Wearable technology has evolved into a low-cost continuous measuring modality providing accurate measurements of activity, movement and energy expenditure. The technology opens up the potential to more accurately research the behavior of patients in the ictal and interictal phases of headache disorders and holds potential as a digital biomarker for CH attacks.

Our real-world objective actigraphic data challenge the stereotypical idea of behavior during CH attacks in patients using acute treatment. To summarize, our analysis suggests that patients with CCH may show reduced AEE and reduced presence of high-intensity movements not only during the ictal phase but also during the pre-ictal phase of CH attacks.

Sense of restlessness and/or agitation are common symptoms of (untreated) CH attacks, hence their integration in the ICHD-3 diagnostic criteria for CH ^1,24^ . Almost 70% to almost 90% of patients with CH described typical signs of psychomotor agitation (restlessness) during the attack in prospective studies ^25^. A prospective clinical survey study in patients with CH found 93% of participants reporting restlessness during the attacks or that movement did not exacerbate the pain ^26^. Another prospective survey study found that 88.1% of participants with CH exhibited signs of psychomotor agitation (restlessness) with an inability to keep still or performing different types of actions during typical and untreated attacks ^25^. Previous scientific reports on the clinical features also documented the sense of restlessness or even compulsed movement as a prominent feature of CH ^5,24,27–29^ Restlessness or agitation as an associated symptom may even be triggered with calcitonin gene-related peptide or nitroglycerin in laboratory-induced CH attacks ^30,31^. Therefore, restlessness is a highly sensitive and highly specific parameter for CH ^24^. Of note that multiple studies in Asia (including in Taiwan, Japan and Korea) have reported on lower percentages of feelings of restlessness and uncoupling from restless behavior ^32–35^.

We infer multiple reasons for our findings of pre-ictal and ictal hypoactivity which, in turn, create new hypotheses and questions for future research. First, based on the data, the behavioral effects of ictal treatment could force the patient into hypoactivity. This seems especially true for oxygen-treated attacks in our dataset, which can be observed in Figure 3 and Appendix 3. Due to the specific context of the wrist actigraphy and despite being worn on the non-dominant wrist, it is possible that patients keep their hand and wrist in a more fixed position, for example to hold the oxygen mask or holding their heads with their hands during the attacks. It can be speculated that these actions might limit intense hands or body movements. It is plausible that chest-worn or hip-worn actigraphy devices yield different results due to the difference between axial and appendicular motions. Second, while the ICHD-3 criteria require severe to very severe pain for untreated attacks, patients in our study reported the severity of the CH attack as moderate, which may affect behavior (see Table 2). In our study, no restrictions were imposed on acute treatment. Hence, the capability to manage attacks, and the high treatment efficacy, could account for both the moderate attack intensity and reduction in AEE. Third, from analyzing the literature, the results may not be completely surprising since previous research efforts may have hinted at reduced AEE in relation to certain CH attacks. Snoer et al. analyzed the presence of symptoms in the pre-ictal, ictal and postictal phase of CH attacks in a prospective, observational questionnaire-based study. Apart from 54% CH attacks having restlessness as a general symptom, also 30.4% CH attacks were accompanied by decreased energy levels ^2^. The authors also found that the most frequent general symptom in the pre-ictal phase was a sense of restlessness (22.2% of attacks), but also that 13.2% of attacks had decreased energy levels and 5.6% of attacks had fatigue as a pre-ictal symptom ^2^. Snoer et al. have also provided good evidence that general symptoms within the pre-ictal phase of CH attacks occurred at a median of 20 minutes prior to the attack, in line with our findings of the most significant drop in AEE between 30 minutes before onset and onset. Furthermore, this Danish study also found “decreased energy levels” and/or “fatigue” as possible pre-ictal and ictal symptoms in their cohort. Furthermore, for the pre-ictal phase, Blau and Engel already reported in 1998 that several patients may feel “tired”, “low”, “apathetic”, “listless”, “withdrawn”, “quiet” or “ill” in the pre-ictal phase ^36^.

We have no doubt that the pain from CH attacks can indeed be excruciating, may lead to a sense of restlessness or agitation, and therefore result in a tendency to pace or move intensely in contrast to the ictal behavior of patients during migraine attacks. This has been documented for many years by patients, clinical experts and researchers. We are however convinced of the quality of our measured data points, the registrations in the headache applications performed by participants, and the data analysis. Our data however suggests that, in real-world settings with no restrictions imposed upon proven highly efficacious acute treatment for CH attacks, we have to be careful with the assumption of increased AEE calculated from wrist-worn actigraphy as this may lead to false positives ^37,38^. In the age of rapid access to actigraphy devices, artificial intelligence, and machine learning algorithms, this feature should be addressed carefully when designing data-driven CH detection models for real-world use.

## Limitations

Limitations to our study should be addressed. First, the low number of patients in our study and the low number of attacks registered make it hard to draw definitive conclusions. The results are preliminary and part of a larger research effort into the detection of behavioral changes of headache patients before, during and after attacks. Second, our study design utilized streaming data, potentially resulting in fewer headaches fulfilling the headache data ratio requirement due to missing data. Böttcher et al. observed that using the Empatica in a non-streaming mode generally yields a higher data ratio, but streaming results in a lower user burden as patients do not have to manually download the data from the device and send it to the cloud ^19^. Third, we cannot extrapolate our findings immediately to patients with ECH since only attacks from patients with longstanding CCH were investigated. Fourth, the study of untreated, and thus more severe attacks, most likely yields different data during the headache phase of the CH attack, and preventive treatment may have its influence too. Our real-world observational study design allowed patients to use all types of CH treatments, both acute and preventive, at their discretion as it was deemed unethical to restrict the use of highly effective treatments at this stage of the research.

## Conclusion

Our findings show that wrist-worn actigraphy for the detection of CH attacks in ambulatory environments with no restrictions on acute treatment may measure reduced activity during attacks, in contrast to our initial assumption that patients would show increased activity energy expenditure due to movement during attacks. Our data may provide a methodological and hypothesis-generating basis for future real-world actigraphy studies in a larger sample of patients with CH. Further research is required to determine the use of wrist-worn actigraphy for the analysis of movement during CH attacks and as a digital biomarker for CH.

## Author Contributions

Study concept and design: all authors. Acquisition of data: Nicolas Vandenbussche, Koen Paemeleire. Analysis and interpretation of data: Nicolas Vandenbussche, Jonas Van Der Donckt. Drafting of the manuscript: Nicolas Vandenbussche, Jonas Van Der Donckt. Revising it for intellectual content: all authors. Final approval of the completed manuscript: all authors.

## Conflicts of Interest

Nicolas Vandenbussche has received travel grants and consulting fees from Novartis AG, TEVA Pharmaceuticals Industries Ltd., AbbVie/Allergan and Pfizer Inc. Koen Paemeleire has received personal compensation from AbbVie/Allergan, Amgen/Novartis AG, Eli Lilly and Company, Lundbeck, Pfizer, Teva Pharmaceuticals Industries Ltd., and Man&Science for consulting, serving on a scientific advisory board, and/or speaking; he and is/was a clinical trial investigator for Almirall (almotriptan), Amgen/Novartis AG (erenumab), Eli Lilly and Company (galcanezumab, lasmiditan), Lundbeck (eptinezumab) and Autonomic Technologies Inc. (sphenopalatine ganglion stimulation). Jonas Van Der Donckt, Mathias De Brouwer, Bram Steenwinckel, Marija Stojchevska, Femke Ongena and Sofie Van Hoecke report no conflicts of interest.

## Funding

Nicolas Vandenbussche is funded by Ghent University Hospital “Fund for Innovation and Clinical Research (Fonds voor Innovatie en Klinisch Onderzoek) 2019” as a PhD Fellow. Jonas Van Der Donckt (1S56322N) is funded by a doctoral fellowship of the Research Foundation Flanders (FWO). Part of this work is done in the scope of the imec.AAA Context-aware health monitoring project.

## Abbreviations

AEE: Activity Energy Expenditure, AI^ABS^ : absolute activity index, CH : cluster headache, CCH: chronic cluster headache, ECH: episodic cluster headache, G: gravitational force, H: variance window length, ICHD-3: International Classification of Headache Disorders, Third Edition, N : number, SD: standard deviation, T: time -point, σ_m_ = signal variance of wrist acceleration over axis x, y or z

## Data Availability

The data is not available.

## Acknowledgements

The authors would like to thank all participants of the study for their volunteering.

## Clinical Trials Registration Number

NCT04949204 (www.ClinicalTrials.gov).

## Appendix 1

**Table:** Descriptive analysis of headache duration.

| description | N | mean+/-std | min | 25% | 50% | 75% | max |
| --- | --- | --- | --- | --- | --- | --- | --- |
| All headaches | 34 | 35min+/-22min | 8min | 20min | 30min | 47min | 1h42min |
| Nighttime headaches | 13 | 32min+/-23min | 9min | 15min | 29min | 42min | 1h25min |
| Daytime headaches | 21 | 37min+/-23min | 8min | 21min | 31min | 50min | 1h42min |
| Daytime headaches<br>w/ >=75% data during | 16 | 38min+/-25min | 8min | 21min | 31min | 50min | 1h42min |
| Daytime headaches<br>w/ >=75% data during<br>& eligible non CH<br>data | 15 | 38min+/-26min | 8min | 20min | 31min | 51min | 1h42min |

## Appendix 2

**Table:**
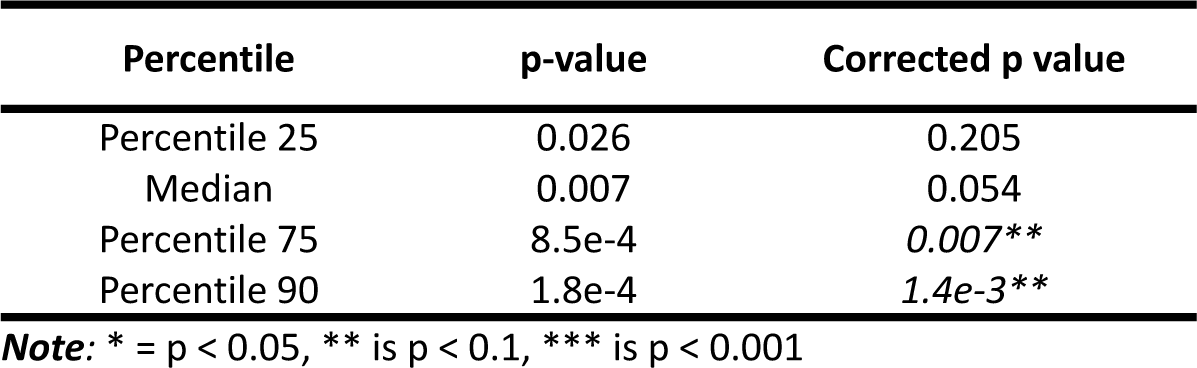
Accompanying p-values of whole headache ΔAI^ABS^ (N=15, Figure 3 - all AI^ABS^ pairs).

**Appendix 2:** **Table:** Accompanying p-values of whole headache $\Delta AI^{ABS}$ (N=15, Figure 3 - all $AI^{ABS}$ pairs).
| Percentile | p-value | Corrected p value |
| --- | --- | --- |
| Percentile 25 | 0.026 | 0.205 |
| Median | 0.007 | 0.054 |
| Percentile 75 | 8.5e-4 | 0.007** |
| Percentile 90 | 1.8e-4 | 1.4e-3** |

**Note:** \* = $p < 0.05$ , \*\* is $p < 0.1$ , \*\*\* is $p < 0.001$

## Appendix 3

**Table:** Accompanying p-values of whole headache ΔAI^ABS^ for oxygen-acute-treated CH attacks (N=9, Figure 3 - oxygen treatment group).

| Percentile | p-value | Corrected p value |
| --- | --- | --- |
| Percentile 25 | 0.004 | 0.031* |
| Median | 0.004 | 0.031* |
| Percentile 75 | 0.004 | 0.031* |
| Percentile 90 | 0.004 | 0.032* |

**Note:** \* = $p < 0.05$ , \*\* is $p < 0.1$ , \*\*\* is $p < 0.001$

## Appendix 4

**Table:** Accompanying p-values of ΔAI^ABS^ intervals relative to headache onset (Figure 4).

**Appendix 4:****Table: Accompanying p-values of $\Delta AI^{ABS}$ intervals relative to headache onset (Figure 4).**
| Time interval | Percentile | p-value | Corrected p-value |
| --- | --- | --- | --- |
| -3h to -1h<br>(N=11) | Percentile 25 | 0.123 | 1 |
|  | Median | 0.123 | 1 |
|  | Percentile 75 | 0.206 | 1 |
|  | Percentile 90 | 0.413 | 1 |
| -1h to -30min<br>(N=12) | Percentile 25 | 1 | 1 |
|  | Median | 0.791 | 1 |
|  | Percentile 75 | 0.129 | 1 |
|  | Percentile 90 | 0.021 | 0.672 |
| -30min to onset<br>(N=11) | Percentile 25 | 0.464 | 1 |
|  | Median | 0.083 | 1 |
|  | Percentile 75 | 0.002 | 0.063 |
|  | Percentile 90 | 0.005 | 0.156 |
| Onset to 30 min<br>(N=14) | Percentile 25 | 0.194 | 1 |
|  | Median | 0.024 | 0.785 |
|  | Percentile 75 | 0.007 | 0.215 |
|  | Percentile 90 | 0.003 | 0.098 |
| 30 min to 1h<br>(N=13) | Percentile 25 | 0.068 | 1 |
|  | Median | 0.032 | 1 |
|  | Percentile 75 | 0.010 | 0.336 |
|  | Percentile 90 | 0.005 | 0.148 |
| 1h to 2h<br>(N=10) | Percentile 25 | 0.322 | 1 |
|  | Median | 0.275 | 1 |
|  | Percentile 75 | 0.160 | 1 |
|  | Percentile 90 | 0.064 | 1 |
| 2 to 3h<br>(N=10) | Percentile 25 | 0.131 | 1 |
|  | Median | 0.193 | 1 |
|  | Percentile 75 | 0.131 | 1 |
|  | Percentile 90 | 0.048 | 1 |

**Note:** \* = $p < 0.05$ , \*\* is $p < 0.1$ , \*\*\* is $p < 0.001$

## Appendix 5

**Figure:**
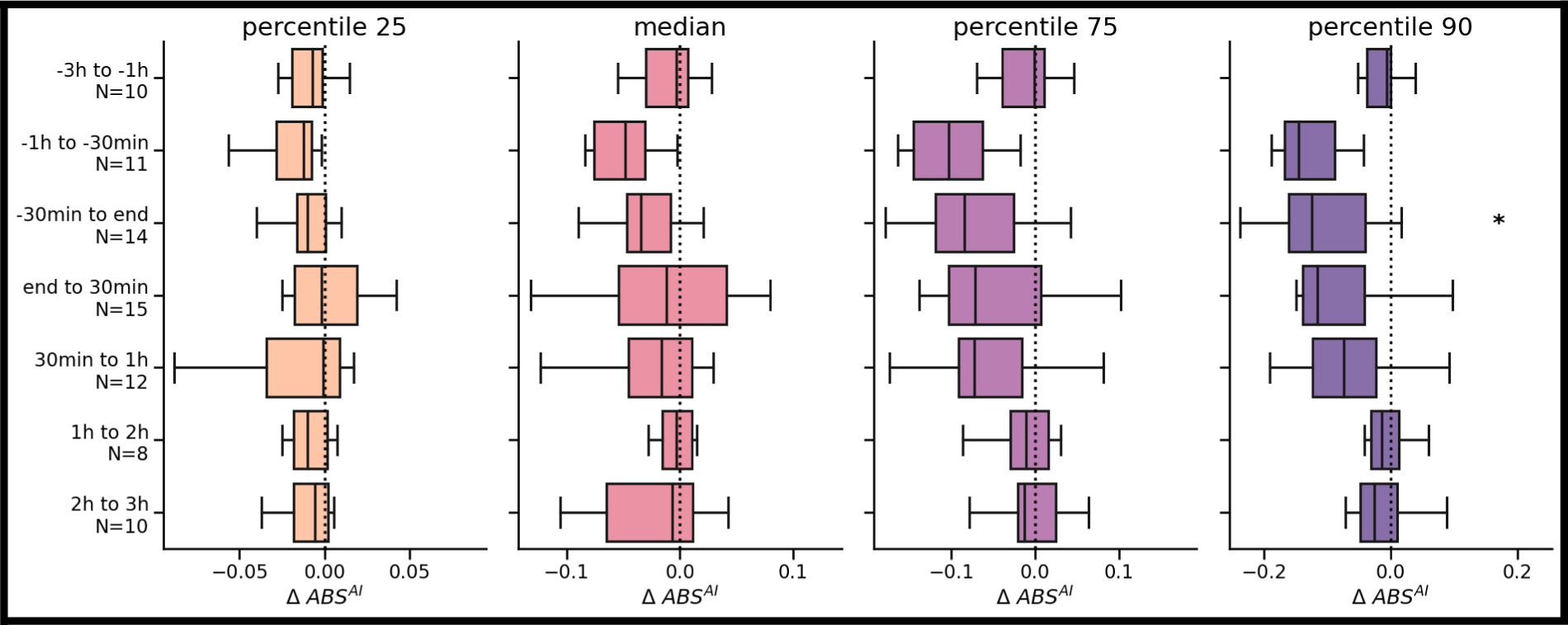
Headache intervals relative to CH end. The y-axis labels indicate the number of attacks considered, which varies due to the >=75% data ratio requirement for both CH and non-CH intervals. *Abbreviations*: AI^ABS^ = absolute activity index, ΔAI^ABS^ = difference between absolute activity indices, CH = cluster headache, h = hour, N = number. Levels of statistical significance (after Bonferroni correction for multiple testing): * p < 0.05; ** p < 0.01; *** p < 0.001.

## Notes

### Author Declarations

Ethics committee/IRB of Ghent University Hospital gave ethical approval for this work

